# Analysis of twenty-week time-series of confirmed cases of New Coronavirus COVID-19 and their simple short-term prediction for Georgia and Neighboring Countries (Armenia, Azerbaijan, Turkey, Russia) in amid of a global pandemic

**DOI:** 10.1101/2020.09.09.20191494

**Authors:** Avtandil G. Amiranashvili, Ketevan R. Khazaradze, Nino D. Japaridze

## Abstract

Results of a comparative statistical analysis of the daily data associated with New coronavirus COVID-19 infection of confirmed cases (Č) of the population in Georgia (GEO), Armenia (ARM), Azerbaijan (AZE), Turkey (TUR) and Russia (RUS) amid a global pandemic (WLD) in the period from March 14 to July 31, 2020 are presented.

The analysis of data is carried out with the use of the standard statistical analysis methods of random events and methods of mathematical statistics for the non-accidental time-series of observations.

In particular, a correlation and autocorrelation analysis of the observational data was carried out, the periodicity in the time- series of Č were revealed, the calculation of the interval prediction values of Č taking into account the periodicity in the time-series of observations from August 1 to 31, 2020 (ARM, AZE) and from August 1 to September 11, 2020 (WLD, GEO, TUR, RUS) were carried out.

Comparison of real and calculated predictions data on Č in the study sites from August 1 to August 31, 2020 is carried out. It was found that daily, monthly and mean weekly real values of Č for all the studied locations practically fall into the 99% confidence interval of the predicted values of Č for the specified time period.

A dangerous situation with the spread of coronavirus infection may arise when the mean weekly values of Č of the 99% upper level of the forecast confidence interval are exceeded within 1–2 weeks. Favorable – when the mean weekly values of Č decrease below 99% of the lower level of the forecast confidence interval.

## Introduction

The New coronavirus (COVID-19) epidemic began in China at the end of 2019 and spread rapidly around the world. On March 11, 2020, a pandemic of this virus was recognized [1].

Despite the taken measures, in general, the growth of morbidity and mortality continues. Traditionally, the main burden in the fight against the pandemic falls on the doctors-epidemiologists. Together with them, scientists and specialists of various disciplines from different countries of the world joined in intensive research of an unprecedented phenomenon (including Georgia and neighboring countries - Armenia, Azerbaijan, Turkey, Russia [2-5]).

For example, experts from the field of physical and mathematical sciences together with doctors make an important contribution to research on the spread of the new coronavirus COVID-19. Despite the fact that scientists are dealing with extremely unstable time series of observations, depending on many factors (medical, methodology for identifying infected, natural, economic, social, political, emotional, religious, mental, etc.), which greatly complicates their systematization and forecasting, they proposed a variety of mathematical and statistical spatial-temporary models of the spread of the new coronavirus [6-17].

Georgia takes the fight against coronavirus seriously, closely monitoring the trends of the pandemic’s variability in the world and especially in its neighboring countries (Armenia, Azerbaijan, Turkey, Russia). In particular, in the paper [5] the preliminary results of a comparative statistical analysis of the mean weekly data associated with new coronavirus infection of confirmed, recovered and fatal cases in the population of the above-mentioned countries amid a global pandemic in the period from March 14 to July 31, 2020 are presented.

This work is a continuation of the above research [5]. This paper presents a detailed statistical analysis of data on daily values of confirmed cases of coronavirus infection in Georgia and neighboring countries in the context of a global pandemic, and also considers the possibility of using one of the simple statistical models to predict the variability of this infection, taking into account the periodicity of its changeability.

We used standard methods of statistical analysis of random events and methods of mathematical statistics for non-random time series of observations.

## Study areas, Material and Methods

The study areas: World (WLD), Georgia (GEO), Armenia (ARM), Azerbaijan (AZE), Turkey (TUR) and Russia (RUS).

Data of John Hopkins COVID-19 Time Series Historical Data (with US State and County data) [https://www.soothsawyer.com/john-hopkins-time-series-data-with-us-state-and-county-city-detail-historical/] about daily values of confirmed coronavirus-related cases, from March 14 to August 21, 2020 are used.

In the proposed work the analysis of data is carried out with the use of the standard statistical analysis methods of random events and methods of mathematical statistics for the non-accidental time-series of observations [18-19].

The following designations will be used below: Mean – average values; Min – minimal values; Max – maximal values; Range – Max-Min; St Dev – standard deviation; σ_m_ – standard error; C_V_ = 100·St Dev/Mean – coefficient of variation, %; Median – median value; R^2^– coefficient of determination; R – coefficient of linear correlation; R_K_ – Kendall rank correlation coefficient; R_S_ – Spearman rank correlation coefficient; R_a_ – coefficient of autocorrelation, Lag = 1 Day; K_DW_ – Durbin-Watson statistic; Res – residual components (RC); Calc – calculated data; Real – measured data from March 14 to July 31 for statistical analysis (twenty weeks, 140 days); Real_1 – measured data for predictions calculation (84 or 42 days); Real_2 – measured data for comparison with calculated predictions data (from August 1 to August 31, 2020); 99%(+/−) – 99% confidence interval of average (the calculations were carried out without taking into account autocorrelation in the time-series of observations); 99%_Low – 99% lower level of confidence interval; 99%_Upp – 99% upper level of confidence interval; Δ = 100· (1-Parameter/Real_2),% - relative difference between measured and calculated values, (forecast relative error, %); α – the level of significance; Daily Values of Confirmed Coronavirus-Related Cases – Č.

The statistical programs Data Fit 7, Mesosaur and Excel 16 were used for calculations.

The curve of trend is equation of the regression of the connection of the investigated parameter with the time at the significant value of the determination coefficient and such values of K_DW_, where the residual values are accidental. If the residual values are not accidental the connection of the investigated parameter with the time we will consider as simply regression.

The calculation of the interval prognostic values of Č taking into account the periodicity in the time-series of observations was carried out using Excel 16.

The following rule of thumb for interpreting the size of a correlation coefficient (CC) is used [20]: 0 ≤ CC < 0.3 – Negligible correlation, 0.3 ≤ CC < 0.5 – Low correlation, 0.5 ≤ CC < 0.7 – Moderate correlation, 0.7 ≤ CC < 0.9 – High correlation, 0.9 ≤ CC ≤ 1.0 – Very high correlation.

## Results and Discussion

The results in the Fig. 1-8 and Table 1-7 are presented.

In Fig. 1 and Table 1 data about statistical characteristics of daily values of confirmed corona-virusrelated cases, from March 14 to July 31, 2020 in Georgia and neighboring countries (Armenia, Azerbaijan, Turkey, Russia) amid a global pandemic are presented.

In particular, analysis of this table reveals the following:
Over the entire observation period, the largest number of infected per 1 million populations was on average observed in Armenia (93.1, range of change 2.0–260.7), the smallest – in Georgia (2.2, range of change 0–11.3). In the World, these values are respectively 16.2, 1.4 and 43.8.
The largest variations in Č values are observed in Azerbaijan (C_V_ = 86.8%), the smallest in Russia (C_V_ = 55.8%). Worldwide C_V_ = 54.9%.

Dynamics of time variability of the values of Č is as follows:

World. Continuous growth, high (R, R_S_) and very high (R_K_) positive correlation with time.
Georgia. Small variability, negligible (R, R_K_, R_S_) negative correlation with time.
Armenia. On the whole moderate (R_K_) and high (R, R_S_) positive correlation with time.
Azerbaijan. Generally, very high (R_S_) and high (R, R_S_) positive correlation with time.
Turkey. Low (R, R_K_, R_S_) negative correlation with time.
Russia. Moderate (R), low (R_S_) and negligible (R_K_) positive correlation with time.

**Fig. 1.**
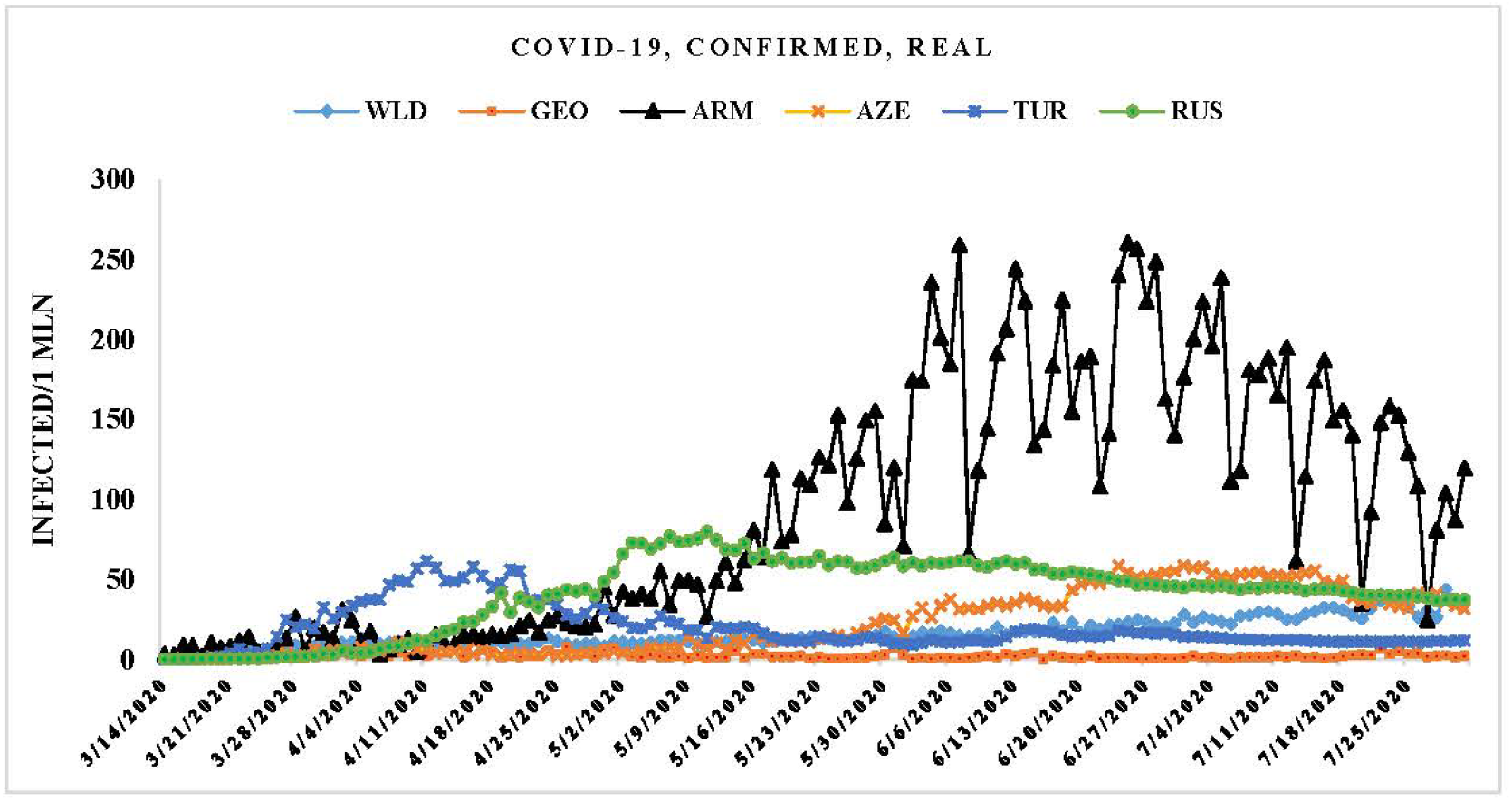
Changeability of the real daily values of new coronavirus-related confirmed cases from March 14 to July 31, 2020 in the study sites.

**Table 1.** Statistical characteristics of daily values of confirmed new coronavirus-related cases, from March 14 to July 31, 2020 in the study sites (per 1 million population)

| Location | World | Georgia | Armenia | Azerbaijan | Turkey | Russia |
| --- | --- | --- | --- | --- | --- | --- |
| Mean | 16.2 | 2.2 | 93.1 | 22.7 | 19.8 | 41.1 |
| Min | 1.4 | 0.0 | 2.0 | 0.0 | 0.0 | 0.0 |
| Max | 43.8 | 11.3 | 260.7 | 58.7 | 61.6 | 79.9 |
| Range | 42.4 | 11.3 | 258.6 | 58.7 | 61.6 | 79.9 |
| Median | 12.9 | 1.6 | 75.7 | 13.7 | 14.3 | 45.0 |
| St Dev | 8.9 | 1.8 | 77.6 | 19.7 | 13.9 | 22.9 |
| $\sigma_m$ | 0.7 | 0.2 | 6.6 | 1.7 | 1.2 | 1.9 |
| $C_v$ (%) | 54.9 | 82.9 | 83.4 | 86.8 | 70.5 | 55.8 |
| Correlation with days |  |  |  |  |  |  |
| $R^2$ ($\alpha \leq 0.08$) | 0.856 | 0.022 | 0.572 | 0.788 | 0.151 | 0.304 |
| $R$ ($\alpha \leq 0.08$) | 0.93 | -0.15 | 0.76 | 0.89 | -0.39 | 0.55 |
| $R_k$ ($\alpha \leq 0.22$) | 0.84 | -0.07 | 0.63 | 0.73 | -0.32 | 0.17 |
| $R_s$ ($\alpha \leq 0.16$) | 0.96 | -0.12 | 0.81 | 0.91 | -0.36 | 0.40 |

The time dependence of measured values of Č have fairly complicated behavior (Table 2, Fig. 2).

**Table 2.** Form of the equations of the regression of the time changeability of the daily values of new coronavirus-related confirmed cases from March 14 to July 31, 2020 in the study sites. The level of significance of R^2^ is not worse than 0.001.

| Location | World | Georgia | Armenia |
| --- | --- | --- | --- |
| Regression | Fourth Order Polynomial | Seven Order Polynomial | Seven Order Polynomial |
| $R^2$ | 0.943 | 0.338 | 0.830 |
| $K_{DW}$ | 1.65 | 1.78 | 1.55 |
| Autocorrelation of RC | Absent | Absent | Absent |
| Location | Azerbaijan | Turkey | Russia |
| Regression | Six Order Polynomial | Tenth Order Polynomial | $\check{C} = \exp(a+b/X+c \cdot \log(x))$ |
| $R^2$ | 0.972 | 0.942 | 0.958 |
| $K_{DW}$ | 0.89 | 0.96 | 0.39 |
| Autocorrelation of RC | Positive | Positive | Positive |

World – fourth order polynomial; Georgia and Armenia **-** seven order polynomial; Azerbaijan **-** six order polynomial; Turkey – tenth order polynomial; Russia **-** Č = exp(a+b/X+c·log(x)), where x are days.

At the same time, autocorrelation of residual components of Č for World, Georgia and Armenia is absent, and for Azerbaijan, Turkey and Russia it is positive. That is, for the first three locations, the calculated curve of dependence Č from time can be considered a trend, for the second three locations – a regression.

**Fig. 2.**
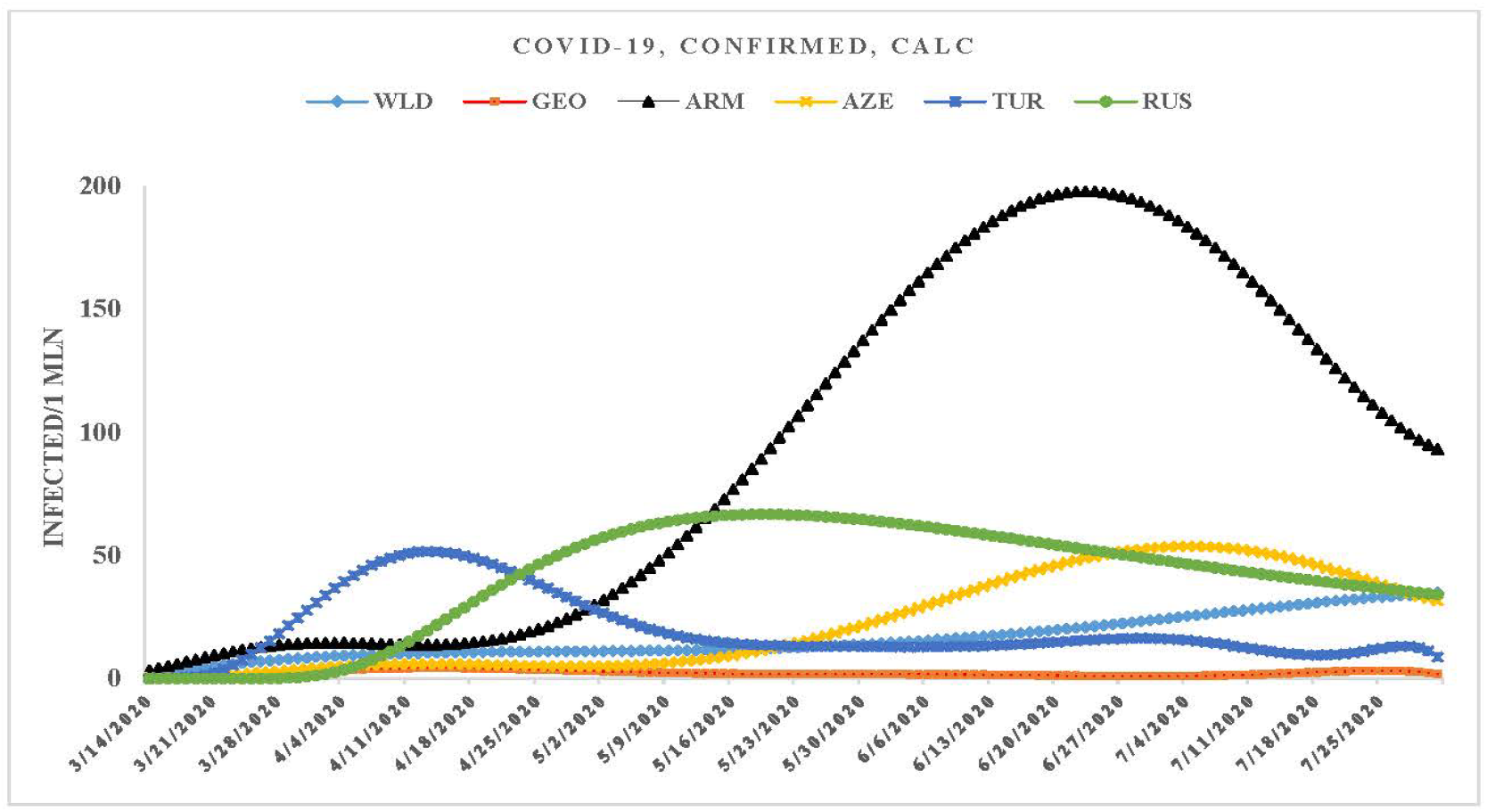
Changeability of the calculated daily values of new coronavirus-related confirmed cases from March 14 to July 31, 2020 in the study sites.

The smoothed out curves of variability of calculated values of Č have the following features (Fig. 2).

World. Continuous growth, min on 3/14/2020 (0.9 infected per 1 mln population), max on 7/31/2020 (34.7 infected per 1 mln population).

Georgia. Small variability in time with two periodic bursts: 4/12/2020–4/16/2020 (4.3 infected per 1 mln population) and 7/23/2020–7/26/2020 (3.1 infected per 1 mln population)

Armenia. Unimodal distribution with right skewness. The peak falls on 6/23/2020 of the study period (197.8 infected per 1 mln population). Then – decrease.

Azerbaijan. Growth up to 7/4/2020–7/5/2020 of the studied observation period (53.7 infected per 1 mln population), then – decrease.

Turkey. Unimodal distribution with left asymmetry. The peak falls on 4/13/2020 of the studied observation period (51.6 infected per 1 mln population). Then – a more or less monotonous decline with a slight surges.

Russia. A vaguely pronounced peak on 5/19/2020 (66.7 infected per 1 mln population) with a further monotonous decrease.

Thus, the nature of the temporal variability of Č values for the studied locations differs significantly from each other. This indicates that the spread of coronavirus infection in these countries during the study period was isolated in nature, associated with the closure of borders and the practically cessation of the movement of people from country to country.

This fact is clearly demonstrated by the data in Table 3. As follows from this table, there is practically no linear correlation between the studied sites by random components of Č values (with the exception of the Armenia-World pair, – Low correlation). The linear correlation between the investigated sites in terms of real Č values basically is significant. However, this correlation is most likely indirect in nature and indicates a similarity or difference in the development trends of a new coronavirus infection in these countries, which was noted in [5]. Otherwise, there would be a correlation for the random components of Č values.

**Table 3.** Linear correlation between the study sites on the daily values of confirmed new coronavirus-related cases from March 14 to July 31, 2020 (the right side of the matrix – real data, the left side – residual components; R min = 0.135, α = 0.1; R max = 0.83, α = < 0.005).

| Location | World | Georgia | Armenia | Azerbaijan | Turkey | Russia |
| --- | --- | --- | --- | --- | --- | --- |
| World | 1 | -0.06 | 0.63 | 0.82 | -0.27 | 0.30 |
| Georgia | 0.04 | 1 | -0.31 | -0.28 | 0.46 | -0.11 |
| Armenia | 0.30 | -0.03 | 1 | 0.83 | -0.42 | 0.51 |
| Azerbaijan | 0.05 | -0.12 | 0.08 | 1 | -0.39 | 0.36 |
| Turkey | 0.06 | -0.06 | 0.02 | -0.02 | 1 | -0.21 |
| Russia | -0.03 | -0.08 | -0.04 | 0.11 | 0.10 | 1 |

The degree of expandability of coronavirus infection can be judged using autocorrelation analysis of a time-series of observations of Č. As follows from Table 4, in all studied locations (with the exception of Georgia), there is a fairly strong autocorrelation in the Č time-series (connectivity in the series, the influence of previous events on subsequent events).

**Table 4.** Number and range of coefficients of autocorrelation and values of peaks of periodicity of the daily values of new coronavirus-related confirmed cases in the study sites.

| Location | Range of Autocorrelation, Lag, Day | Range of Coefficient of Autocorrelation, $\alpha \leq 0.05$ | Peak of Periodicity, Day |
| --- | --- | --- | --- |
| World | 1÷11 | 0.92÷0.62 | 7 |
| Georgia | 1÷4; 14 | 0.41 ÷ 0.28; 0.33 | 14 |
| Armenia | 1÷14 | 0.86÷0.72 | 7 |
| Azerbaijan | 1÷13 | 0.98÷0.79 | 28 |
| Turkey | 1÷9 | 0.96÷0.63 | 20 |
| Russia | 1÷11 | 0.98÷0.71 | 20 |

World – in the first 11 lags, variability – from very high to moderate autocorrelation.

Georgia – in the first 4 lags and 14^th^ lag (low – negligible values of R_a_).

Armenia – in the first 14 lags, variability – within high autocorrelation.

Azerbaijan – in the first 13 lags (very high – high values of R_a_).

Turkey – in the first 9 lags, variability – within very high to high autocorrelation.

Russia – in the first 11 lags, variability – from very high to moderate autocorrelation.

Note, that R_a_ values close to 1 in the first lag could determine the presence of a positive autocorrelation of the residual components of the Č time-series in Azerbaijan, Turkey, and Russia (Table 2).

In table 4 data on values of peaks of periodicity of the C in the study sites are presented also (World, Armenia – 7 days; Georgia – 14 days; Azerbaijan – 28 days; Turkey, Russia – 20 days).

As follows from Fig. 1,2 and Table 2 in the period from 3/14/2020 to 31/7/2020 changeability in time of the daily values of new coronavirus-related confirmed cases in all locations have a nonlinear nature. Therefore, for the interval forecast of the Č values, we selected time-series with a linear tendency of variability of the daily values of new coronavirus-related confirmed cases after their peak values. The forecast period is half of the previous time-series of the Č for World, Georgia, Turkey and Russia, and about 75% – for Armenia and Azerbaijan (Table 5).

**Table 5.**
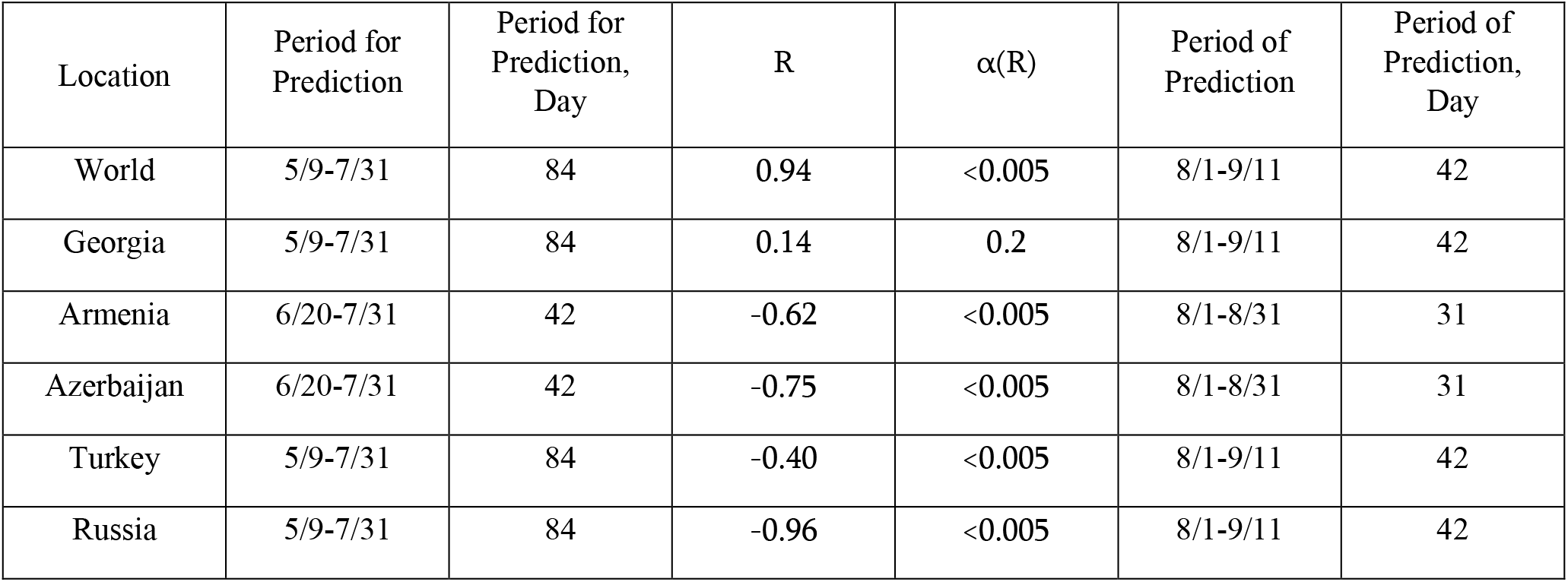
Periods for prediction and of prediction of the daily values of new coronavirus-related confirmed cases in the study sites.

In Fig. 3-8 the results of calculating the forecast of the values of Č for the studied locations (taking into account on the periodicity in the time-series of observations) and their 99% of the upper and lower confidence intervals from August 1 to 31, 2020 are shown. For comparison with the calculated values, the same figures show data on the real values of Č.

**Fig. 3.**
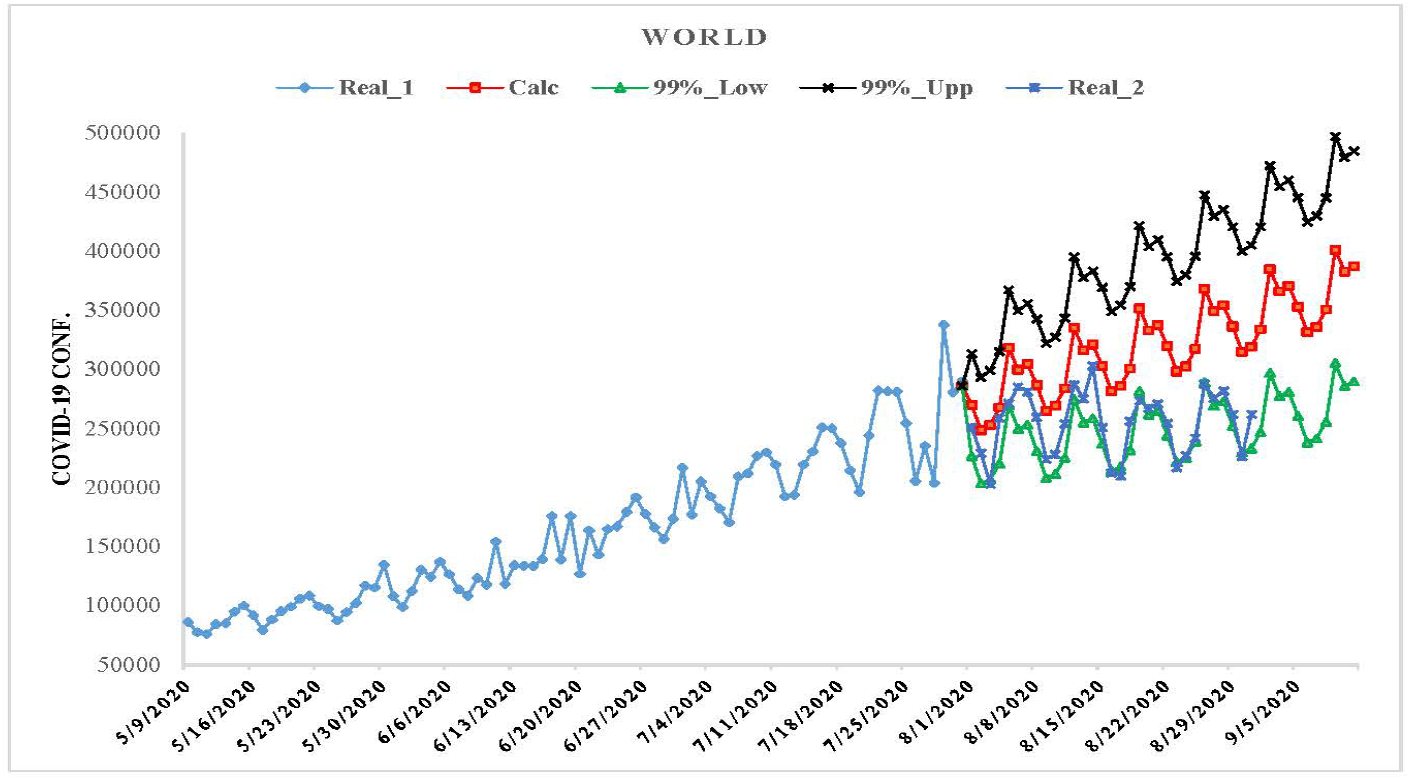
Six-week prediction of the daily values of new coronavirus-related confirmed cases for world

**Fig. 4.**
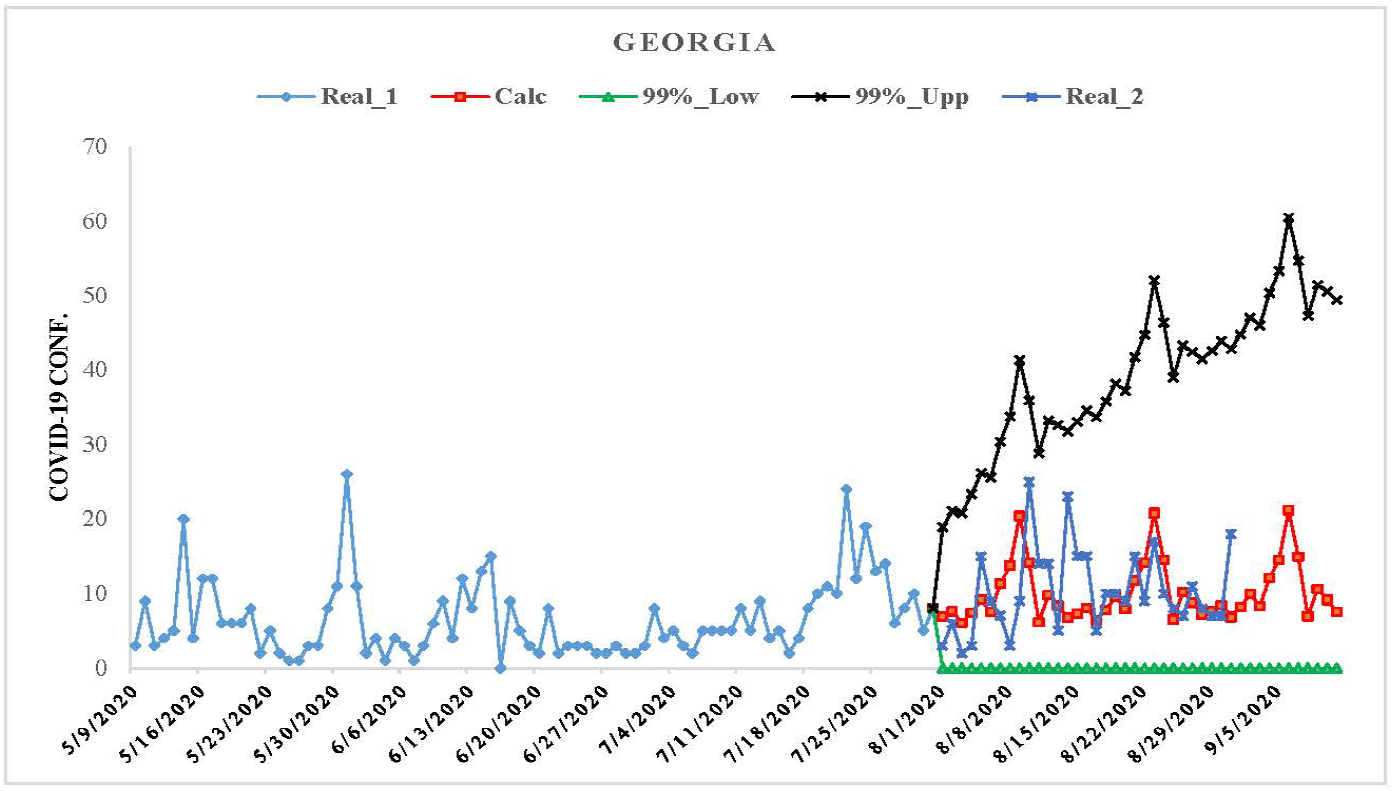
Six-week prediction of the daily values of new coronavirus-related confirmed cases for Georgia

**Fig. 5.**
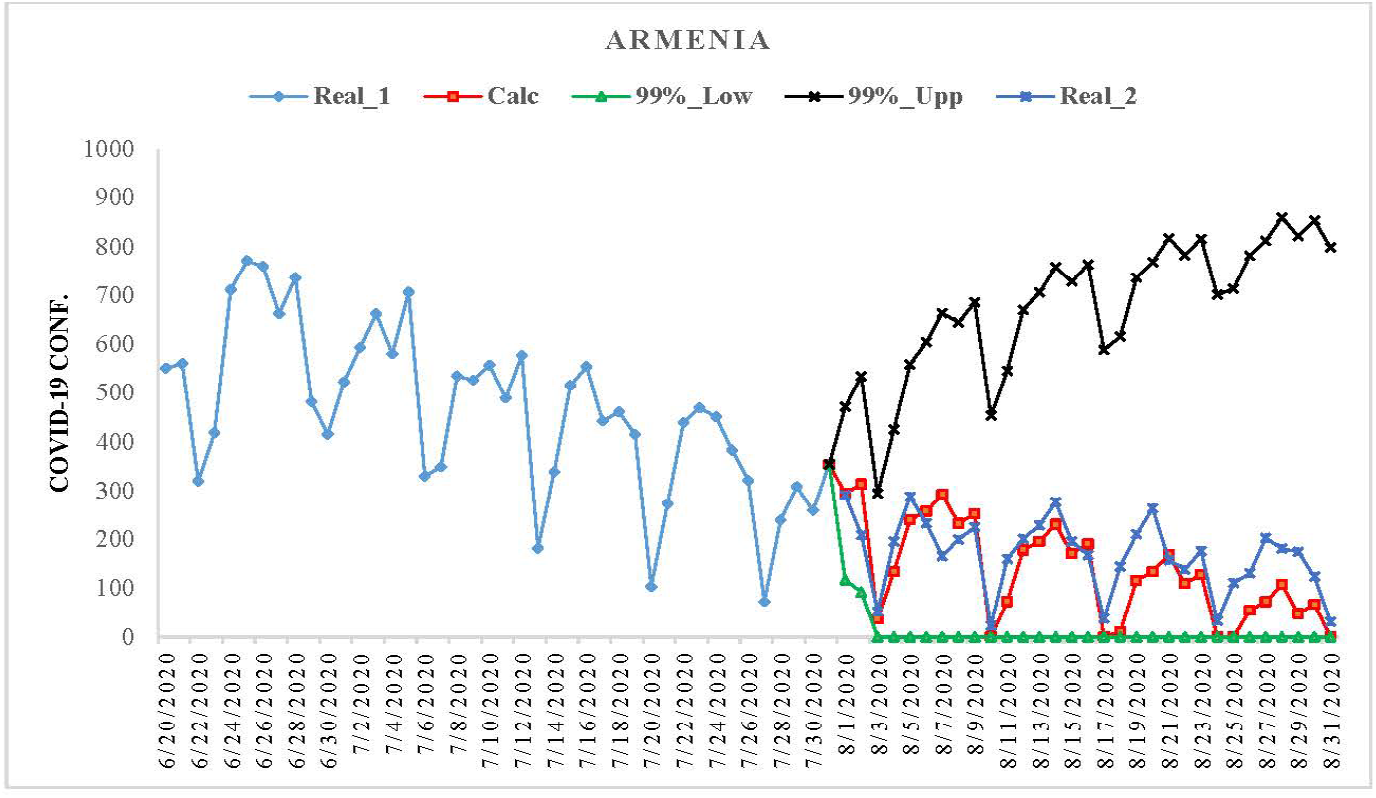
Monthly prediction of the daily values of new coronavirus-related confirmed cases for Armenia

**Fig. 6.**
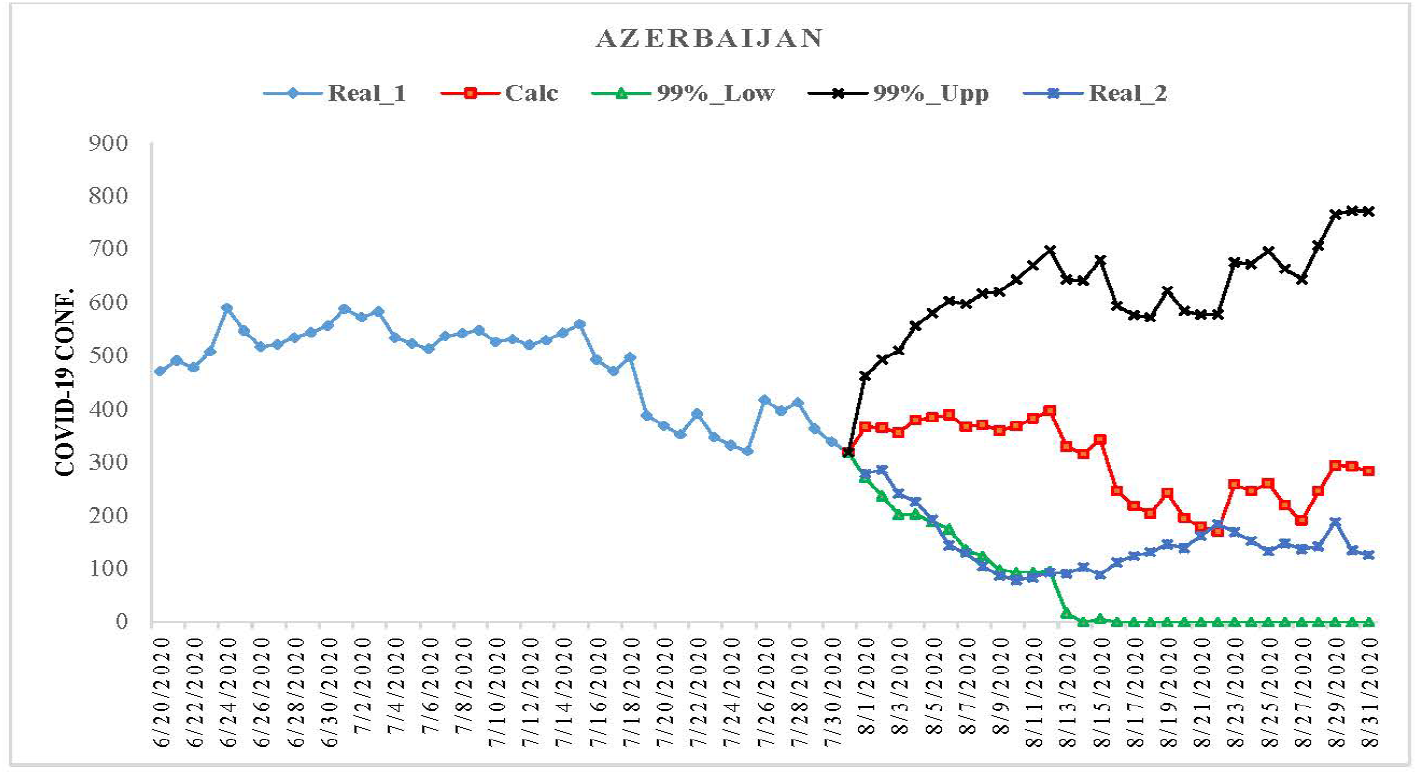
Monthly prediction of the daily values of new coronavirus-related confirmed cases for Azerbaijan

**Fig. 7.**
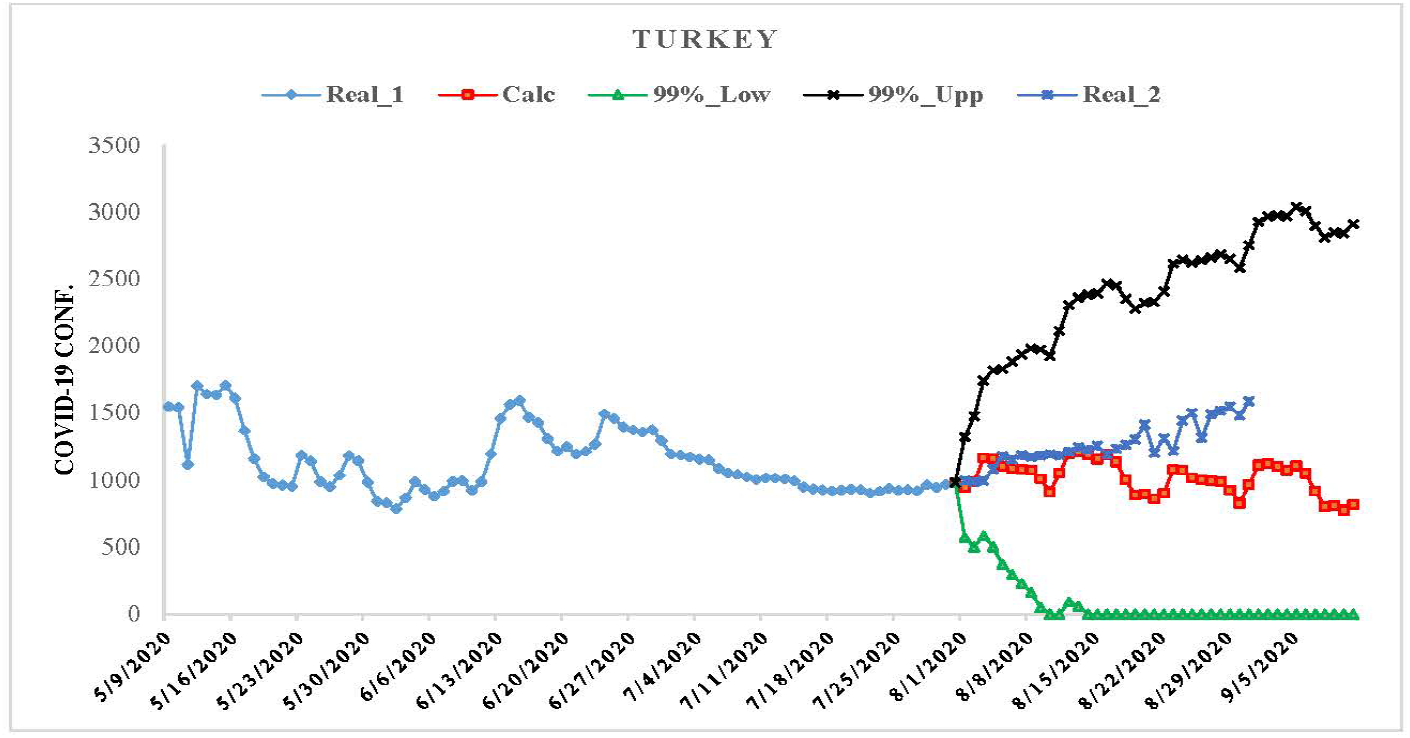
Six-week prediction of the daily values of new coronavirus-related confirmed cases for Turkey

**Fig. 8.**
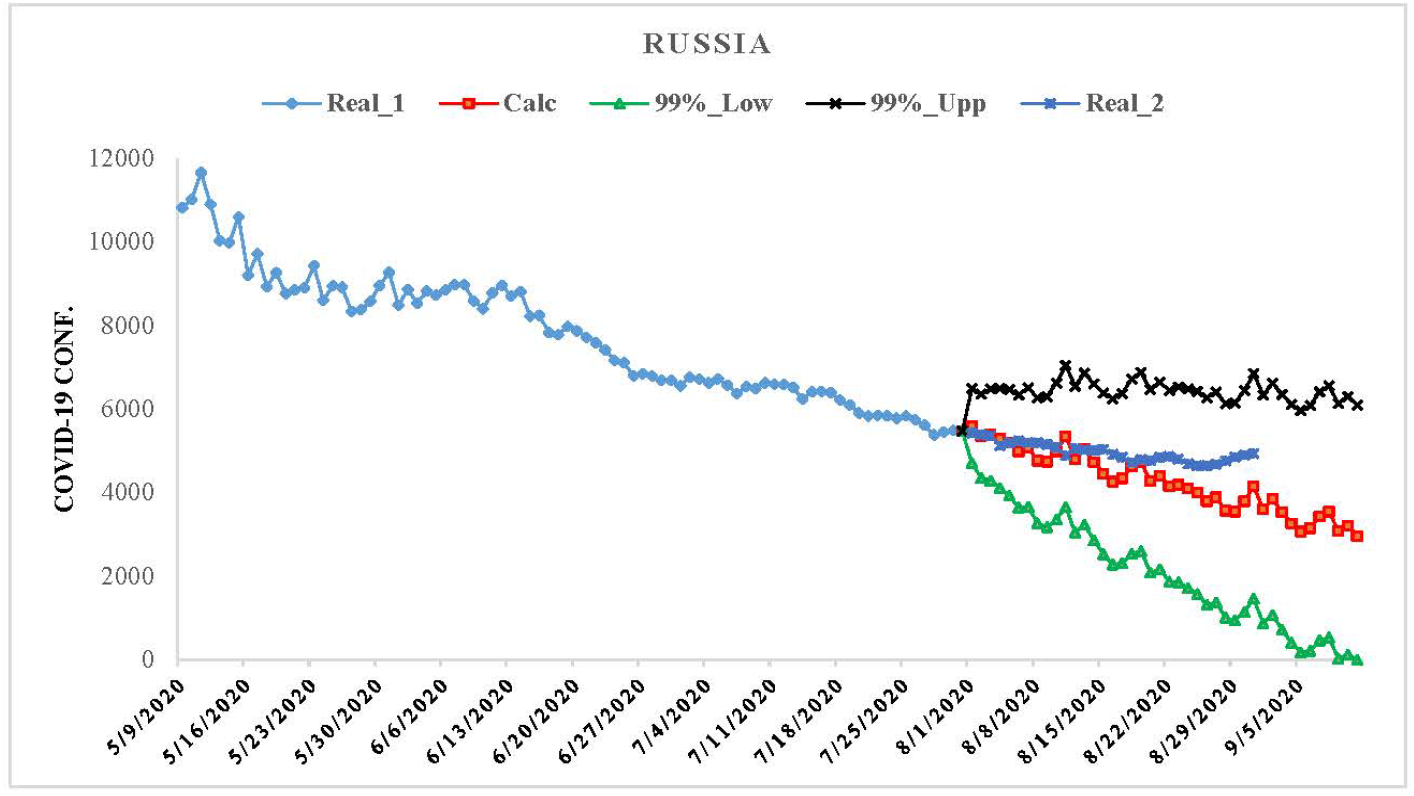
Six-week prediction of the daily values of new coronavirus-related confirmed cases for Russia

As follows from Fig. 3-8 and tables 6,7 daily, monthly and average weekly real values of Č for all the studied locations practically fall into the 99% confidence interval of the predicted values of Č for the specified time period.

Tables 6 and 7 also provide data on the coincidence of real and forecasted monthly and weekly average values of Č.

The best match between the calculated and real monthly mean values of Č (values of Δ) is as follows.

World: 5% with 99%_Low level; Georgia: 6% with calculated values within the confidence interval; Armenia: 21% with calculated values within the confidence interval; Azerbaijan: 57% with 99%_Low level; Turkey: 18% with calculated values within the confidence interval; Russia: 8% with calculated values within the confidence interval (Table 6).

**Table 6.** Comparison of mean values of real and calculated predictions data of the daily values of new coronavirus-related confirmed cases in the study sites from August 1 to August 31, 2020.

| Parameter | Calc | 99%_Low | 99%_Upp | Real_2 | Calc | 99%_Low | 99%_Upp | Real_2 |
| --- | --- | --- | --- | --- | --- | --- | --- | --- |
| Location | World |  |  |  | Georgia |  |  |  |
| Max | 368238 | 288979 | 447496 | 303009 | 21 | 0 | 52 | 25 |
| Min | 248524 | 203724 | 293324 | 202692 | 6 | 0 | 19 | 2 |
| Range | 119714 | 85255 | 154172 | 100317 | 15 | 0 | 33 | 23 |
| Mean | 306876 | 241295 | 372457 | 254353 | 10 | 0 | 35 | 10 |
| St Dev | 30862 | 24014 | 40866 | 26395 | 4 | 0 | 8 | 6 |
| 100·(1-Par/Real_2),% | -21 | 5 | -46 |  | 6 | 100 | -244 |  |
| Location | Armenia |  |  |  | Azerbaijan |  |  |  |
| Max | 314 | 118 | 859 | 291 | 397 | 272 | 772 | 286 |
| Min | 0 | 0 | 294 | 23 | 170 | 0 | 462 | 79 |
| Range | 314 | 118 | 565 | 268 | 227 | 272 | 310 | 207 |
| Mean | 133 | 7 | 676 | 169 | 297 | 63 | 628 | 147 |
| St Dev | 99 | 27 | 139 | 74 | 74 | 87 | 75 | 54 |
| 100·(1-Par/Real_2),% | 21 | 96 | -301 |  | -102 | 57 | -328 |  |
| Location | Turkey |  |  |  | Russia |  |  |  |
| Max | 1208 | 584 | 2752 | 1587 | 5599 | 4703 | 7039 | 5429 |
| Min | 826 | 0 | 1321 | 987 | 3544 | 939 | 6128 | 4639 |
| Range | 382 | 584 | 1431 | 600 | 2055 | 3764 | 911 | 790 |
| Mean | 1034 | 110 | 2254 | 1266 | 4566 | 2644 | 6488 | 4966 |
| St Dev | 109 | 193 | 376 | 162 | 574 | 1080 | 212 | 225 |
| 100·(1-Par/Real_2),% | 18 | 91 | -78 |  | 8 | 47 | -31 |  |

The best match between the calculated and real weekly mean values of Č Č (range of absolute values of Δ) is as follows.

World: from 8 to 1 % with 99%_Low level; Georgia: from 15 to 26 % with calculated values within the confidence interval; Armenia: from 9 to 51 % with calculated values within the confidence interval; Azerbaijan: from 6 to 19 % with 99%_Low level and from 80 to 49 % with calculated values within the confidence interval; Turkey: from 1 to 28 % with calculated values within the confidence interval; Russia: from 0 to 16 % with calculated values within the confidence interval (Table 7).

**Table 7.** Comparison of weekly mean values of real and calculated predictions data of the daily values of new coronavirus-related confirmed cases in the study sites from August 1 to August 28, 2020.

| Location | Weekly Mean Parameters |  |  |  |  | 100·(1-Par/Real_2),% |  |  |
| --- | --- | --- | --- | --- | --- | --- | --- | --- |
|  | Period | Calc | 99%_Low | 99%_Upp | Real_2 | Calc | 99%_Low | 99%_Upp |
| WLD | 8/1-8/7 | 280219 | 232839 | 327599 | 254029 | -10 | 8 | -29 |
|  | 8/8-8/14 | 296805 | 237749 | 355861 | 261567 | -13 | 9 | -36 |
|  | 8/15-8/21 | 313391 | 244222 | 382559 | 248604 | -26 | 2 | -54 |
|  | 8/22-8/28 | 329976 | 251638 | 408315 | 255031 | -29 | 1 | -60 |
| GEO | 8/1-8/7 | 8 | 0 | 24 | 6 | -25 | 100 | -269 |
|  | 8/8-8/14 | 11 | 0 | 34 | 13 | 15 | 100 | -155 |
|  | 8/15-8/21 | 8 | 0 | 36 | 11 | 26 | 100 | -222 |
|  | 8/22-8/28 | 12 | 0 | 44 | 10 | -17 | 100 | -342 |
| ARM | 8/1-8/7 | 224 | 30 | 507 | 205 | -9 | 85 | -147 |
|  | 8/8-8/14 | 166 | 0 | 637 | 188 | 12 | 100 | -239 |
|  | 8/15-8/21 | 113 | 0 | 717 | 168 | 33 | 100 | -327 |
|  | 8/22-8/28 | 68 | 0 | 781 | 139 | 51 | 100 | -462 |
| AZE | 8/1-8/7 | 372 | 202 | 543 | 214 | -74 | 6 | -154 |
|  | 8/8-8/14 | 360 | 75 | 648 | 92 | -292 | 19 | -604 |
|  | 8/15-8/21 | 232 | 1 | 601 | 129 | -80 | 99 | -366 |
|  | 8/22-8/28 | 227 | 0 | 662 | 152 | -49 | 100 | -336 |
| TUR | 8/1-8/7 | 1076 | 436 | 1715 | 1082 | 1 | 60 | -59 |
|  | 8/8-8/14 | 1091 | 51 | 2148 | 1202 | 9 | 96 | -79 |
|  | 8/15-8/21 | 1018 | 0 | 2370 | 1266 | 20 | 100 | -87 |
|  | 8/22-8/28 | 1008 | 0 | 2609 | 1399 | 28 | 100 | -87 |
| RUS | 8/1-8/7 | 5269 | 4093 | 6446 | 5274 | 0 | 22 | -22 |
|  | 8/8-8/14 | 4913 | 3224 | 6602 | 5057 | 3 | 36 | -31 |
|  | 8/15-8/21 | 4442 | 2357 | 6527 | 4842 | 8 | 51 | -35 |
|  | 8/22-8/28 | 3955 | 1529 | 6381 | 4723 | 16 | 68 | -35 |

It should be noted that the period of the best forecast of the weekly mean values of Č within the confidence interval is different for different locations. World: 2 weeks (abs Δ = 10–13%); Georgia: 2 weeks (abs Δ = 25–15%); Armenia: 2 weeks (abs Δ = 9–12%); Azerbaijan: – low accuracy within the confidence interval; Turkey: 3 weeks (abs Δ = 1–20%); Russia: 4 weeks (abs Δ = 0–16%).

It should also be noted that, in our opinion, a dangerous situation with the spread of coronavirus infection may arise when the mean weekly values of Č of the 99% upper level of the forecast confidence interval are exceeded within 1–2 weeks. Favorable – when the mean weekly values of Č decrease below 99% of the lower level of the forecast confidence interval.

It also should be paid attention to the increase or decrease in the degree of autocorrelation in the time-series of observations.

## Conclusion

It is planned to continue similar studies for Georgia and neighboring countries in the future, in particular, clarifying data on the periodicity of processes, checking the temporal representativeness of short-term interval statistical forecasting of the new coronavirus infection COVID-19, taking into account the data on periodicity, etc.

## Data Availability

https://www.soothsawyer.com/john-hopkins-time-series-data-with-us-state-and-county-city-detail-historical/

## References

1. World Health Organization. (2020). Coronavirus disease 2019 (COVID-19). Situation report. 67.

2. Meister S, Eradze I, Grigoryan A, Samadov B. The COVID-19 Pandemic in the South Caucasus. ETH Zurich Research Collection. Available from: https://doi.org/10.3929/ethz-b-000415805

3. Öztoprak F, Javed A. (2020). Case fatality rate estimation of COVID-19 for European countries: Turkey’s current scenario amidst a global pandemic. Comparison of outbreaks with European countries. EJMO. 4(2): 149–159, DOI: 10.14744/ejmo.2020.60998.

4. Zemtsov SP, Baburin VL. (2020). Risks of morbidity and mortality during the COVID-19 pandemic in Russian regions. Population and economics. 4(2): 158–181. Available from: https://doi.org/10.3897/popecon.4.e54055

5. Amiranashvili AG, Khazaradze KR, Japaridze ND. (2020). Twenty Weeks of the Pandemic of Coronavirus Covid-19 in Georgia and Neighboring Countries (Armenia, Azerbaijan, Turkey, Russia). Preliminary Comparative Statistical Data Analysis. International Scientific Conference „Modern Problems of Ecology“, Proceedings, ISSN 1512–1976. Tbilisi-Telavi, Georgia, 26–28 September, 2020; 7: 364–370. Available from: https://www.researchgate.net/publication/343945549_Twenty_Weeks_of_the_Pandemic_of_Coronavirus_Covid19_in_Georgia_and_Neighboring_Countries_Armenia_Azerbaijan_Turkey_Russia_Preliminary_Comparative_Statistical_Data_Analysis

6. Ankarali H, Ankarali S, Caskurlu H, Cag Y, Ferhat F, Erdem H, Vahaboglu H. (2020). A Statistical Modeling of the Course of COVID-19 (SARS-CoV-2) Outbreak: A Comparative Analysis. Asia Pacific Journal of Public Health. 32(4): 157–160. Available from: https://journals.sagepub.com/doi/10.1177/1010539520928180

7. Catal’a M, Alonso S, Alvarez-Lacalle E, López D, Cardona P-J, Prats C. Empiric model for short-time prediction of COVID-19 Spreading. medRxiv preprint doi: https://doi.org/10.1101/2020.05.13.20101329.

8. Nishiura H, Linton NM, Akhmetzhanov AR. (2020). Serial interval of novel coronavirus (COVID-19) infections. International Journal of Infectious Diseases. 93: 284–286. Available from: https://doi.org/10.1016/j.ijid.2020.02.060

9. Anastassopoulou C, Russo L, Tsakris A, Siettos C. (2020). Data-based analysis, modelling an. forecasting of the COVID-19 outbreak. PLoS ONE. 15(3): e0230405. Available from: https://doi.org/10.1371/journal.pone.0230405

10. Jung S, Akhmetzhanov AR, Hayashi K, Linton NM, Yang Y, Yuan B, et al. (2020). Real-time estimation of the risk of death from novel coronavirus (COVID-19) infection: inference using exported cases. J. Clin. Med. 9(2)E: 523. Available from: doi:http://dx.doi.org/10.3390/jcm9020523.

11. Tandon H, Ranjan P, Chakraborty T, Suhag V. Coronavirus (COVID-19): ARIMA based time-series analysis to forecast near future. Available from: https://arxiv.org/ftp/arxiv/papers/2004/2004.07859.pdf

12. Aslan IH, Mahir Demi M, Wise MM, Lenhart S. Modeling COVID-19: Forecasting and analyzing the dynamics of the outbreak in Hubei and Turkey. medRxiv preprint doi: https://doi.org/10.1101/2020.04.11.20061952.

13. Ozdinc M, Senel K, Ozturkcan S., Akgul A. Predicting the Progress of COVID-19: The Case for Turkey. Turkiye Klinikleri Journal of Medical Sciences. April 2020. DOI: 10.5336/medsci.2020-75741

14. Batista M. Estimation of a state of Corona 19 epidemic in August 2020 by multistage logistic model: a case of EU, USA, and World. medRxiv preprint doi: https://doi.org/10.1101/2020.08.31.20185165.

15. Petropoulos F, Makridakis S. (2020). Forecasting the novel coronavirus COVID-19. Plos one. 15(3): e0231236.

16. Roosa K, Lee Y, Luo R, Kirpich A, Rothenberg R, Hyman J, et al. (2020). Real-time forecasts of the COVID-19 epidemic in China from February 5th to February 24th, 2020. Infectious Disease Modelling. 5: 256–263.

17. Roosa K, Lee Y, Luo R, Kirpich A, Rothenberg R, Hyman JM, et al. (2020). Short-term forecasts of the COVID-19 epidemic in Guangdong and Zhejiang, China: February 13–23, 2020. Journal of Clinical Medicine. 9(2): 596.

18. Förster E., Rönz B. (1979). Methoden der korrelations – und regressionsanalyse. – Ein Leitfaden für Ökonomen. Verlag Die Wirtshaft Berlin. 324.

19. Kendall MG. (1981). Time-series. Moscow, 200, (in Russian).

20. Hinkle DE., Wiersma W, Jurs SG. (2003). Applied Statistics for the Behavioral Sciences. Boston, MA, Houghton Miffli. Company.

